# Phenome-wide association network demonstrates close connection with individual disease trajectories from the HUNT study

**DOI:** 10.1101/2022.07.18.22277775

**Authors:** Martina Hall, Marit K. Skinderhaug, Eivind Almaas

## Abstract

**Background:** Disease networks offer a potential road map of connections between diseases. Several studies have created disease networks where diseases are connected either based on shared genes or Single Nucleotide Polymorphisms (SNP) associations. However, it is still unclear to which degree SNP-based networks map to empirical co-observed diseases within a different, general, adult study population spanning over a long time period.

**Methods:** We create a SNP-based disease network (PheNet) from a large population using the UK biobank phenome-wide association studies. Importantly, the SNP-associations are adjusted for linkage disequilibrium, case/control imbalances, as well as relatedness. We map the PheNet on to significantly co-occurring diseases in the Norwegian HUNT study population, and further, identify consecutively occurring diseases with significant occurrence in the PheNet.

**Results:** We find that the overlap between the networks are far larger than expected, where most diseases tend to link to diseases of the same category and some categories are more linked to each other than expected by chance. Considering the ordering of consecutively occurring diseases in the HUNT data, we find that many diabetic disorders and cardiovascular disorders are subsequent the diagnostication of obesity and overweight, and cardiovascular disorders that often tend to be observed subsequent to other diseases are associated with higher mortality rates.

**Conclusions:** The HUNT sub-PheNet showing both genetically and co-observed diseases offers an interesting framework to study groups of diseases and examine if they, in fact, are comorbidities and pinpoint exactly which mutation(s) that constitute shared cause of the diseases. This could be of great benefit to both researchers and clinicians studying relationships between diseases.

## Background

Since the first successful Genome-Wide Associations Study (GWAS) in 2007 [1], a large body of scientific work has been devoted to identify candidate genetic loci that influence the risk of developing certain diseases or phenotypes. Currently, the GWAS catalog consist of more than 5,800 publications and 398,000 associations between Single Nucleotide Polymorphisms (SNPs) and multiple diseases [2, 3]. As a result of the need for post-GWAS analyses to interpret the results, phenome-wide association studies (PheWAS) have proven efficient in identifying pleiotropic effects of disease SNPs for a broad range of physiological and/or clinical outcomes based on Electronic Health Records (EHR) [4, 5, 6]. Thus, PheWas analyses offer the opportunity for a system level approach for studying disease interactions.

The network medicine field was initialized by Goh and coworkers when creating a Human Disease Network (HDN) based on causal genes from the Online Mendelian Inheritance in Man (OMIN) database [7]. The HDN represents the linking of diseases that are associated with one or more genes, and it gives a full landscape of known human genetic disorders. In this study, it was discovered that most diseases are actually connected through common genetic origins, as the network consists of a giant component connecting hundreds of diseases in addition to several smaller components [7].

Reusing the successful approach of the HDN, several works have aimed at constructing similar disease networks by instead using detailed SNP-disease connections from GWAS to investigate disease-disease associations at a genomic level [8, 9, 10]. These works were successful at grouping similar diseases based on common genetic SNP findings from GWAS. However, they were based on rather limited sets of diseases (7 to 177) and a limited number of both participants and SNPs analyzed in the GWAS. In addition, as for the HDN study [7], some of these investigations relied on summary statistics from different studies, which could influence the validity of their findings. It is quite possible that differences in phenotype definitions and test-association methods when merging these results into a disease network would impact the outcomes.

A recent study [11] used PheWas summary statistics from a single source EHR, the Geisinger’s biobank, consisting of 625, 325 SNP associations with 541 disease codes from the International Classification of Diseases, Ninth Revision (ICD9). Their constructed disease network consists of 358 diseases linked by 1, 398 connections, showing that many diseases are also genetically linked through common GWAS-significant SNPs [11]. However, their summary statistics originates from a study [12] which uses the PLATO method [13] for association testing. PLATO applies a logistic regression model which does not account for relatedness of the participants or for imbalance in the case/control ratios when testing the associations between SNPs and binary phenotypes. Using EHR data from the UK Biobank (UKBB) participants, Dong and coworkers [14] created a similar GWAS-based disease network and compared to observed comorbidities within the UKBB participants. Their SNP disease associations were based on GWAS data from a linear mixed model [15] for both binary and continuous traits, which also does not account for imbalances in case/control ratios and is suboptimal for binary traits. Note that, comparisons with comorbidities within the same study population introduces bias in terms of evaluating the relationship among the diseases outside of the study population.

A related study using PheWas summary statistics from UKBB was analyzed using SAIGE (Scalable and Accurate Implementation of GEneralized mixed model) [16, 17]. In association testing with SAIGE, the sample relatedness and unbalanced case/control ratios are adjusted for. Using a *p*-value threshold of 10^*−*4^ and grouping SNPs into linkage disequilibrium (LD) clusters, the authors created a disease network consisting of 1, 403 phenotypes and focused on identifying comorbidities related to obstetric disorders. They validated the use of disease ego-centric networks by comparing the genetic risk of comorbidity with the neighbouring diseases to empirically observed comorbidities for the same study participants. However, as pointed out by the authors, in order to get a reliable validation of their results, one would need to compare observed comorbidities to data from a different study population [16].

Here, we use the same UKBB based PheWas summary statistics as in Ref. [16], but with adjusteed criteria for inclusions of phenotypes and SNP associations (see Methods for details). The aim is to validate the findings of genetically linked diseases from the UKBB based disease network with actual co-occurrences from a different study population, the Trndelag Health Study (HUNT) participants. The HUNT-study is one of the longest longitudinal population studies, covering up to *∼* 90% of the adult population in Nord-Trndelag, Norway from 1984 until 2019 [18, 19]. Matching these participants with EHR and the cause of death registry, we have an almost complete health record history of 90, 103 participants diagnosed with one or multiple of the diagnoses from the UKBB PheWas summary, from August 1987 until June 2017. With this data, we have, to our knowledge, the largest time interval for medical history, and we use it to investigate if the genetically linked diseases correspond to actual comorbidities in a different study population.

## Methods

### Datasets

#### UK Biobank PheWas

We create the phenome-wide association network (PheNet) from summary statistics of 1, 403 binary phenotypes from a broad EHR-based PheWas of *∼* 400, 000 White British participants of European ancestry [17, 20]. The phenotypes are represented as phenotype codes (phenocodes) which is a collection of similar ICD billing codes from the EHR, and are classified into 17 disease categories. The summary statistics were generated using SAIGE, which, unlike many other GWAS association tests, handles unbalanced case/control ratios and relatedness with a generalized linear mixed model, controlling for sex, birth year and four principal components (PC1-PC4) [17]. Using a *p*-value threshold of *<* 10^*−*6^ and including only top hits (lowest *p*-value) of the SNP associations in high LD, the UKBB summary statistics consists of 21, 532 SNP associations with 1, 397 phenocodes at a 2-digits level. We neither use no cutoff for minor allele frequencies (MAF) nor number of cases in order to include as many significant SNP-disease associations as possible, and we assume that unrealistic findings will be filtered out when considering the observed co-morbidity status among the HUNT participants.

#### The HUNT study and related health records

The HUNT study is a population based longitudinal study inviting all adult (age *≥* 20) inhabitants of Nord-Trndelag county in Norway to health related questionnaires and clinical measurements in four 11-year time intervals, ranging from 1984 until 2019 [18, 19]. The first study in 1984 (HUNT1) had a participation rate at 89.4% of the inhabitants of Nord-Trndelag. The next rounds of invitations (HUNT2, HUNT3 and HUNT4) expanded the study to include also short interviews, clinical examinations and biological sampling, as well as expanding the sample population to include those aged 13 *−* 19. For the last survey (HUNT4), also inhabitants of the neighboring Sr-Trndelag county were included. The uniqueness of the HUNT study is the high participation rate with the ability to follow a large fraction of the population over a time interval of up to 35 years. As of 2020, the HUNT study consists of a total of 230, 000 participants [21].

Another strength of the HUNT study is the possibility to link the participants to several local, regional, and national health related registries due to the unique Norwegian 11-digit personal identification number [18]. Such registries include, among others, the Medical Birth Register of Norway, the Norwegian Prescription Database, the Cancer Register of Norway, the Norwegian Cause of Death Register, and regional (the Nord-Trndelag Hospital Trust (HNT)) and national (Norway Control and Payment of Health Reimbursement (KUHR)) registers for hospital and general practitioner records.

In this paper, we use data from HUNT1, HUNT2, and HUNT3, and we link the participants to ICD-billing codes from HNT and KUHR and the Norwegian Cause of Death Registry. With this, we have a complete list of diagnoses made at hospital visits (HNT 1987-2017) and at the general practitioner (KUHR 2006-2017) for a total of 90, 103 patients. We are also able to track participants that have died due to the diseases. As we are interested in validating the findings from the UKBB based PheNet, we consider only ICD codes from HUNT that are mapped to phenocodes existing in the PheNet, resulting in 967 of the 1, 397 UKBB phenocodes. The mapping from ICD9 and ICD10 codes were performed using the PheCode Maps [22, 23].

### Construction of the PheNet

When constricting the PheNet, we first group SNPs that are in high LD, as the single top hit SNPs for different diseases can be in high LD and represent the same genetic cause of the disease, without being a mutation at the exact same loci position. Using the *LDmatrix* function from the LDlinkR R-library [24, 25], SNPs that are within *∼* 500 kb and share a high LD (*r*^2^ *≥* 0.8), are defined into LD-blocks. From the updated list of SNP/LD-block disease associations (from here on mentioned as SNP-disease associations), a bipartite network is created, where a link between SNP and disease is present if the corresponding associated *p*-value is *p <* 10^*−*6^. The disease-disease network is further generated, where two diseases are linked if they share one or more SNPs from the bipartite network.

#### Link weights in the PheNet

To obtain a reasonable measure of the link weight between pairs of diseases, we utilize the effect sizes of the SNP-disease association, *β*. The effect sizes measures the log odds ratio for obtaining the disease given the presence of the SNP, and is hence a reasonable measure for the strength of the association between the SNP and the disease. We argue that our method of merging the effect sizes rather than merging *p*-values or counting the number of associated SNPs provides more information regarding the disease-disease associations.

In order to obtain a single link weight between two diseases sharing one or more associated SNPs, we calculate the link weight according to Fig. 1. In this illustration, there are *n* unique SNPs that are significantly associated with both disease 1 and 2. In the first step, the effect sizes linking the diseases though the same SNP, 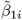 and 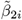, are merged into o<u>ne</u> effect size for SNP *i* by the geometric mean of the absolute effect sizes, 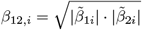. If some of the associated SNPs are in LD, we use the mean effect sizes for these SNPs before calculating the geometric mean. This step results in *n* link weights between disease 1 and 2 when they share associations with *n* common SNPs. Next, to obtain a single link weight between disease 1 and 2 we calculate the arithmetic mean of the *n* link weights between the diseases, such that the final link weight between diseases 1 and 2 is 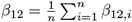.

**Figure 1.**
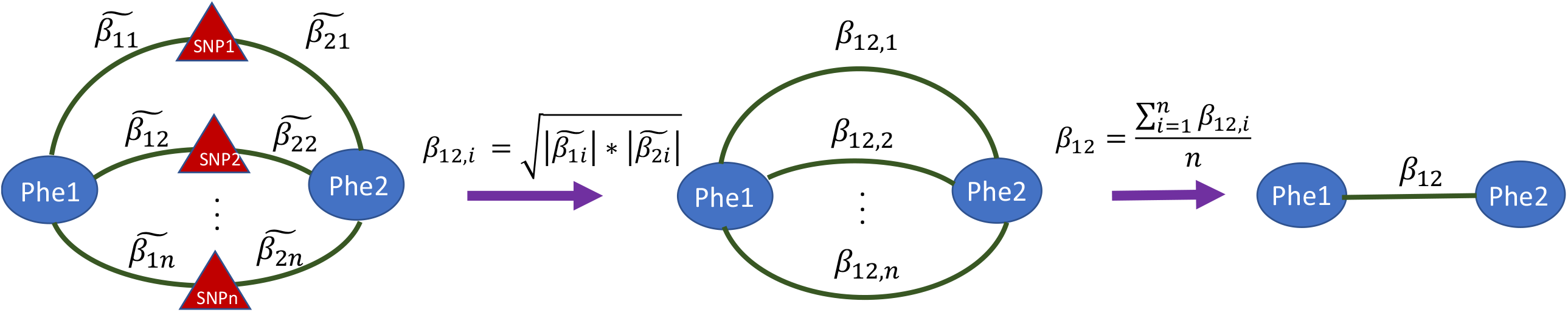
Calculation of link weight for phenotypes sharing common SNPs. The first step involves calculating the geometric mean of the two effect sizes for SNP *i*, 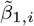 and 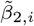 from the UKBB PheWas. In the second step, the *n* common SNPs-effects are combined into one with a arithmetic mean resulting in the final link weight, *β*_1,2_.

### Overlap with co-occurring diseases in the HUNT study

For each HUNT participant, a list of their registered diseases, considering only the 967 phenocode diseases, are ordered based on the first diagnose date. If a person is registered with diseases A, B, C, and D, we construct the pairs A-B, A-C, A-D, B-C, B-D and C-D. Constructing such pairs for all patients, we count the number of times each pair of diseases are present among the 90, 103 participants. To obtain a measure for the strength of co-occurrence for these disease pairs, we use the *ϕ*-score proposed as a comorbidity measure by Hidalgo et. al. [26]. The *ϕ*-score is a Pearson correlation for binary variables, defined as

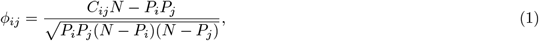

where *N* is the total number of participants, *C*_*ij*_ is the number of patients with disease *i* and *j*, and *P*_*i*_ and *P*_*j*_ is the number of patients with disease *i* and *j* respectively. To assess only the disease pairs with a co-occurrence larger than expected by chance, we perform a one sided *t*-test, with the test statistic defined as

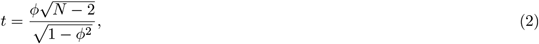

with *N −* 2 degrees of freedom. Extracting only the disease pairs observed in the PheNet, we classify the disease pairs as significantly observed comorbidities if the Bonferroni adjusted *p*-value from the one sided *t*-test is below 0.05*/*1, 135 = 4.405 · 10^*−*5^, where the number 1, 135 is the number of disease pairs tested.

To access the significance of the number of significantly observed disease pairs, we compare our finding with the corresponding finding of random networks, holding the same network properties as the PheNet. Shuffling the labels (disease names) of the nodes, 10^4^ random networks are simulated, and the number of disease pairs with Bonferroni adjusted significant *ϕ*-scores are counted for each network, giving an empirical distribution for the number of significant co-observed pairs in the random networks.

Further, comparing the PheNet disease pairs with the ordered co-occurring disease in the HUNT study population, we perform the same method as above where the disease pairs from the HUNT participants are ordered based on the diagnose date of the disease. A participant with diseases A, B, C and D (in that order) will now give the disease pairs A-B, B-C and C-D. From the constructed frequency list of ordered disease pairs for all participants, the *ϕ*-scores and corresponding *p*-values are calculated. As the PheNet gives no direction of the links, both directions of the disease pairs are considered when extracting these disease pairs from the frequency list of ordered disease pairs and counting the number of pairs with Bonferroni significant *ϕ*-scores. This score is again validated against the empirical distribution of corresponding scores from the 10^4^ random networks.

### Creating the HUNT sub-PheNet

The HUNT sub-PheNet is the sub network of the UKBB based PheNet where only the links with Bonferroni adjusted significant *ϕ*-scores are included. In this way, the HUNT sub-PheNet represents disease associations that are both genetically linked and at the same time being strongly linked as comorbidities based on actual observed disease co-occurrences.

### Network analysis

#### Grouping genetically linked diseases into network modules

As we expect that groups of similar and related diseases will cluster together in the PheNet, we use the Louvain’s network clustering algorithm to construct network modules [27]. This greedy method optimizes the modularity when constructing modules. The modularity measure the density of links within modules compared to links between the modules and is defined as

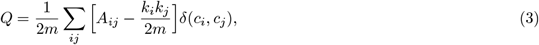

where *A*_*ij*_ represents the link weight between disease *i* and *j, k*_*i*_ = ∑_*j*_ *A*_*ij*_ and *k*_*j*_ = ∑_*i*_ *A*_*ij*_, *m* = ∑_*ij*_ *A*_*ij*_, *c*_*i*_ and *c*_*j*_ are the modules of disease *i* and *j* respectively and *δ* is the Kronecker delta function equal to 1 if *c*_*i*_ = *c*_*j*_ and 0 otherwise.

Using the *cluster louvain* function from the *igraph* R-library [28] we construct modules for both the UKBB based PheNet and the HUNT sub-PheNet using the *β*-scores as link weights for the PheNet and the *ϕ*-score as link weight for the HUNT sub-PheNet. Note that the Louvain’s algorithm does not support negative link weights, but this gives no problems as all *β*-scores are absolute values and all significant *ϕ*-scores are positive in the HUNT sub-PheNet.

#### Disease homogeneity

To test the hypothesis that diseases tend to link to diseases of the same disease category, we create a disease homogeneity score, the *H*-score, representing the diversity of categories linked to a disease [29]. The *H*-score is defined as,

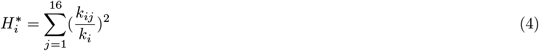

where *k*_*ij*_ is the number of diseases of category *j* linked to disease *i*, and *k*_*i*_ is the degree of disease *i*. This *H*-score is hence very driven by the degree of the disease, as the maximum and minimum value it can take depends on the number of possible categories linked to it. To adjust for this fact, we scale the score such that all *H*-scores take a value between zero and one and are independent of the degree of the disease [30],

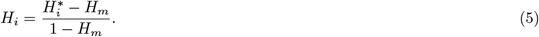

Here, *H*_*m*_ is the minimum value disease *i* can have and is defined according to the degree of the disease, *k*_*i*_,

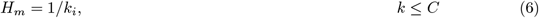

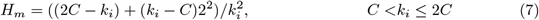

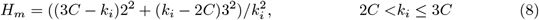

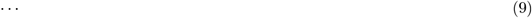

where *C* = 16 is the number of categories in the PheNet. With this, a *H*-score of 1 represents diseases only connected to a single disease category, while a *H*-score of 0 represents maximal difference of categories (two or more categories, and equal amount of diseases from each category).

To test if the mean *H*-score within each module and each category are significantly different from expected, we simulate 10^4^ random networks holding the same properties as the PheNet and the HUNT sub-PheNet. In each of the simulated networks, only the categories are shuffled without replacement, and the distribution of mean *H*-scores within each module and category are used as an empirical distribution to test if the observed corresponding *H*-score from the PheNet and the HUNT sub-PheNet are Bonferroni significantly different from the empirical distribution. The reported *p*-values are the fraction of random mean *H*-scores larger than the observed *H*-scores, Bonferroni adjusted by multiplying with the number of tests (number of modules and number of categories; *n*_PheNet_ = (12, 16) and *n*_*sub−PheNet*_ = (10, 16)).

#### Testing interactions across categories with the Z-score

To test if some categories are more or less linked to each other than what is expected by chance, we calculate a *Z*-score, a normalized score for the number of links shared between each pair of categories,

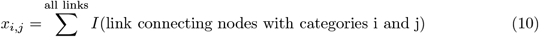

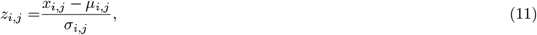

where *µ*_*i,j*_ and *σ*_*i,j*_ are the mean and standard error of *x*_*i,j*_ from 10^4^ simulated random networks. The simulated networks are constructed in the same manner as for the *H*-score analysis, and the significance of the *Z*-score is tested with a twosided *Z*-test from a standard normal distribution. The *p*-values are reported with Bonferroni adjustment considering *n*_PheNet_ = *n*_*sub−PheNet*_ = 16 · 15*/*2 + 16 = 136 tests.

### Direction of disease links and linkage with the Norwegian Cause of Death registry

Finally, we investigate if some of the disease pairs are observed in a specific order, giving a disease history of the HUNT participants. Considering all disease pairs from the PheNet, we test if disease A is more probable to be observed before disease B and vice versa with a binomial test. If the ordering of the diseases are random, the null hypothesis is that drawing A before B has a probability of *p* = 0.5 with *n* being the number of participants with both diseases. For all disease pairs from the PheNet, where the observed number is the number of times disease A are listed before disease B in the HUNT data, we perform a two-sided binomial test and extract only the pairs of diseases with a Bonferroni adjusted *p <* 0.025*/*1, 135 *≈* 2.2 · 10^*−*5^, and further extract only the disease pairs found in the HUNT sub-PheNet. The median time between diseases for all participants with this ordering of co-occurring diseases are registered.

For each of the diseases observed to be in a specific order, termed first disease or last disease, we perform a hypergeometric test to see if some of the disease categories are more or less represented than expected by chance. With a hypergeometric test, we observe *x* of these ordered diseases from *m* available diseases of the specific category, with *n* being the number of trials; the total number of first/last diseases, and *N* being the number of available diseases; the number of diseases in the HUNT sub-PheNet. The *p*-values from this two-sided hypergeometric test of enrichment among the categories are hence calculated as,

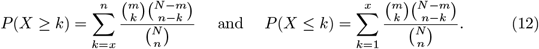

The *p*-values are Bonferroni adjusted with 16 tested categories when reported, where a *p <* 0.025 is considered a significant finding. This test is performed for both sets of first and last diseases separately.

We also link these diseases to the Norwegian Cause of Death registry and register how many who have died from each of these diseases. Note that in the Norwegian Cause of Death registry, diagnoses highly related to the cause of death are also listed. Also, some participants could have their first event of a specific disease as the cause of death and are hence not registered in the hospital nor general practitioner records with this disease.

## Results

### The PheNet shows genomic linkage between diseases

The UKBB based PheNet constructed from the updated list of SNP-disease associations, where all included diseases are available in the HUNT study, includes 457 diseases with 1, 135 links between them. The full PheNet including all diseases from the PheWas summary can be found in Additional file 1, Fig. S1. In the PheNet shown in Fig. 2, most diseases are linked in a giant component and several smaller components, confirming that also at a genomic level, the genetic origin of many diseases are shared with other diseases. The nodes, representing diseases, are colored based on their disease category, sized by the number of associated SNPs to the specific disease and the link between diseases are scaled based on the number of shared SNPs. The twelve largest modules identified by the Louvain’s clustering algorithm with the *β*-scores as link weights are circled in and numbered in the figure. Most diseases are only connected by a few SNPs, such as *Obesity* linked with *Essential hypertension*, while others, such as *Arthropathy NOS* linked with *Other arthropathies* and *Benign neoplasm of uterus* linked with *Uterine leiomyoma* share more than 40 common SNPs. In contrast to other studies [8, 9, 10, 11, 16], this network consists of associations found from a solid framework for genomic association testing even with imbalanced case/control ratios, a large sample population for the association testing (UKBB participants), a stringent threshold for associations (*p*-value *<* 10^*−*6^), and adjustment for LD in linking diseases based on common SNPs.

**Figure 2.**
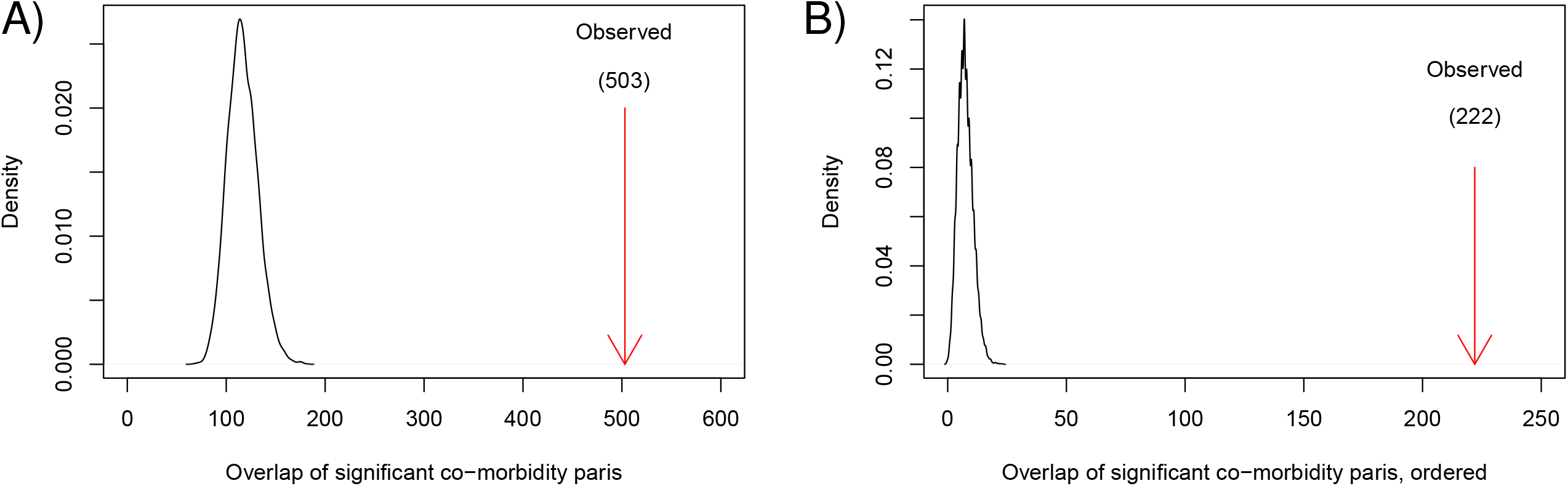
The UKBB based PheNet. The twelve largest modules are marked by circles, the node size corresponds to the number of SNPs associated to the disease and colored based on the disease category. The link thickness corresponds to the link weight between the two diseases. A listing of modules and diseases is given in Additional file 3.

The number of diseases from each category and the mean degree for the disease of each category are shown in Tab. 1. All categories except pregnancy complications are represented, where the neoplasms (*n* = 66) and circulatory system (*n* = 59) are the disease categories with the most diseases. Congenital anomalies (*n* = 5) and infectious diseases (*n* = 5) are the categories with the fewest diseases in the PheNet. Diseases of the circulatory system category have the highest mean degree, on average linked to 8.4 diseases, and most of them are located in module 4, which is dominated by diseases from the circulatory system category (see Additional file 3). This category holds diseases related to cardiovascular diseases, for which many are known to be heritable, and many studies aim to understand the genetic causes of these diseases [31, 32]. Among the links in module 4, we find that *Myocardial infarction, Angina Pectoris* and *Coronary atherosclerosis* are all linked together based on 12, 17 and 19 shared SNPs. These diseases are closely related, as they are all caused by reduced blood flow to the heart, and previous studies have found several genetic markers prone to cause these diseases [33, 34, 35].

**Table 1.**
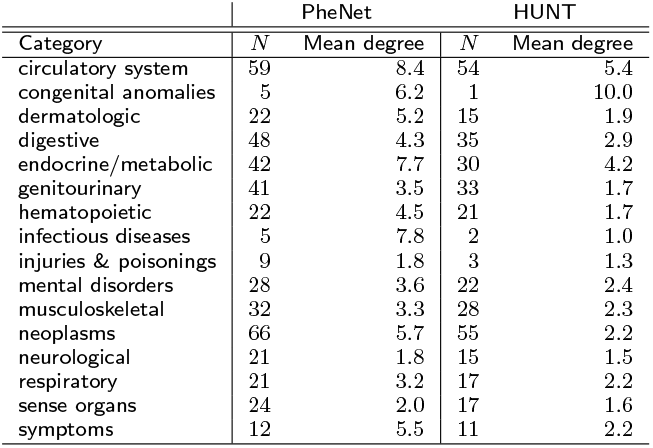
Number of diseases of each category for the PheNet and the HUNT sub-PheNet

### Diseases tend to link within disease categories

Visually inspecting the PheNet in Fig. 2, it seems that diseases from the same category are often linked to one another. This is to be expected, since many of the diseases within a category are quite similar and hence, might share much of the same genetic background. Modules 10 and 12 consist only of diseases from the digestive and neoplasm category respectively, while modules 4 and 7 are dominated by diseases from the circulatory system, mental disorders and neoplasm, with most links connected within the categories. On the other hand, module 2 consists of diseases from many categories and share many links across categories.

To test that the linking within categories are more prominent in the PheNet than expected by chance, we calculate the *H*-score for each disease. The *H*-score represents the diversity of the disease connections, taking into account how many categories each disease is linked to. Diseases with high *H*-scores are “monochromatic” diseases that are connected to mostly the same category, while diseases with low *H*-score are connected to phenocodes of many different categories. In a random network, one would expect the absence of a pattern in regarding the disease connections and hence, observing low *H*-scores. In contrast, in a network where diseases from the same category cluster together, we would expect to find higher *H*-scores for many of the diseases. Fig. 3 A) shows the mean *H*-score within each module, plotted against 10^4^ random networks simulated with the same network properties. We see that for all modules except module 6 and 9, the mean *H*-score in the module is significantly larger than expected (see *p*-values in Tab. 2), supporting our observation that diseases from the same category are more likely to connect to diseases of the same category. Considering the non-significant modules, module 6 is dominated with diseases from the musculoskeletal category where the diseases represent forms of *Arthropathy*, which is diseases of a joint. These diseases are connected to *Diseases of esophagus* of the digestive category in module 2, *Benign neoplasm of uterus* of the neoplasm category in module 7, as well as *Other peripheral nerve disorders, Internal derangement of knee, Unspecified diffuse connective tissue disease* and *Lymphadenitis* of the neurological, injuries and poisoning, dermatological and hematopoietic categories respectively. Module 9 consists of two diseases from the circulatory system (both diseases of *Arterial embolism and thrombosis*) and four diseases from the sense organs category (all four related to *Glaucoma*). These diseases from the module are linked to each other as well as to diseases related to cancer of brain of the neoplasms category in module 7.

**Figure 3.**
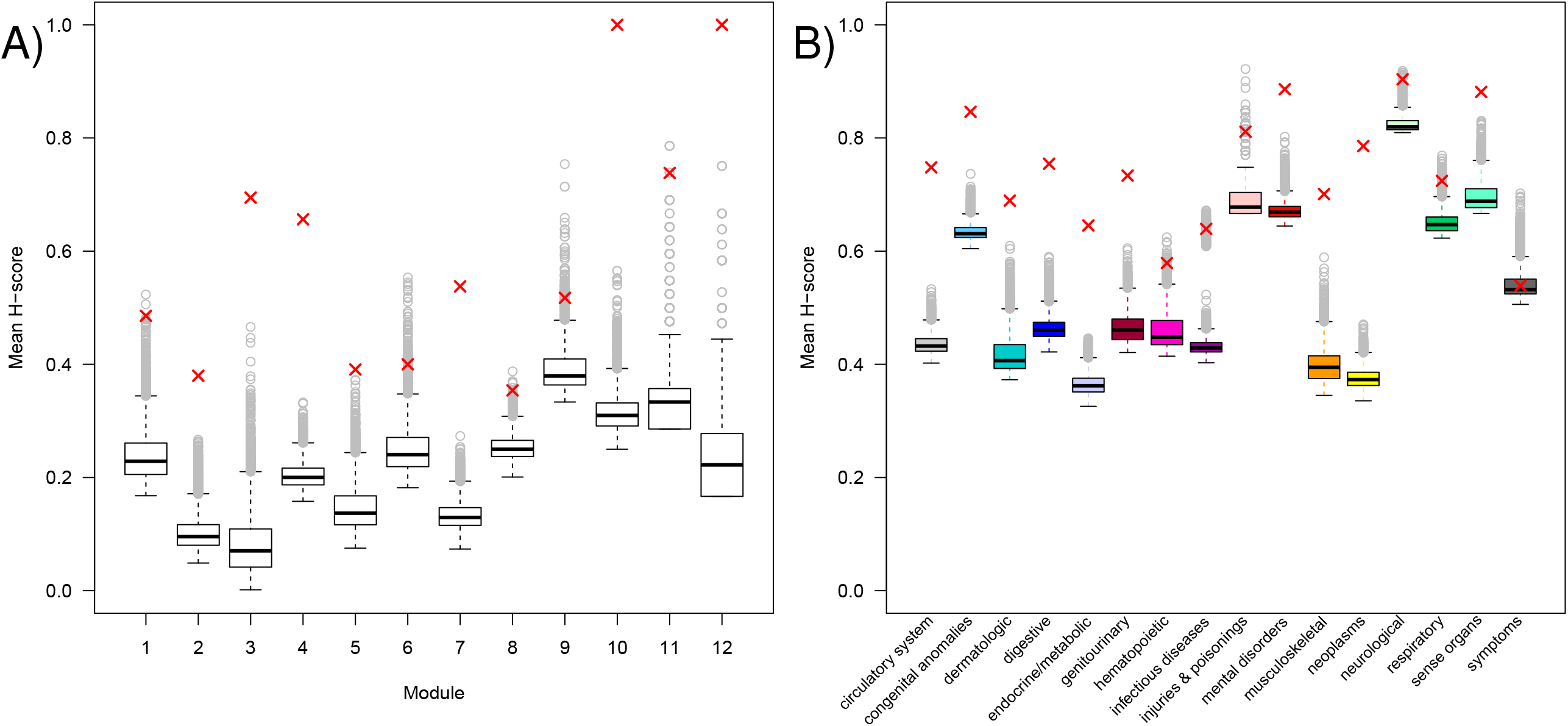
Mean H-score of phenotype network compared to 10^4^ random networks. Mean H score across the twelve largest modules A) and across the 16 phenotype categories B). The red x-es shows the results from the PheNet, while the boxes with whiskers and outliers shows the results from 10^4^ simulated networks.

**Table 2.** Mean H-score with corresponding Bonferroni adjusted p-values for each module.

| PheNet |  |  | HUNT |  |  |
| --- | --- | --- | --- | --- | --- |
| Modules | Mean H | $p$-value | Modules | Mean H | $p$-value |
| 1 | 0.49 | 0.0024 | 1 | 0.51 | 0.02 |
| 2 | 0.38 | $< 10^{-4}$ | 2a | 0.90 | $< 10^{-4}$ |
| | | | 2b | 0.55 | $< 10^{-4}$ |
| 3 | 0.69 | $< 10^{-4}$ | | | |
| 4 | 0.66 | $< 10^{-4}$ | 4a | 0.87 | $< 10^{-4}$ |
| | | | 4b | 0.82 | $< 10^{-4}$ |
| 5 | 0.39 | $< 10^{-4}$ | 5 | 0.86 | $< 10^{-4}$ |
| 6 | 0.40 | 0.1236 | 6 | 0.62 | 0.02 |
| 7 | 0.54 | $< 10^{-4}$ | 7 | 0.66 | $< 10^{-4}$ |
| 8 | 0.35 | 0.0084 | 8 | 0.46 | 0.01 |
| 9 | 0.52 | 0.1524 |  |  |  |
| 10 | 1.00 | $< 10^{-4}$ | 10 | 1.00 | $< 10^{-4}$ |
| 11 | 0.74 | 0.0084 |  |  |  |
| 12 | 1.00 | $< 10^{-4}$ | | | |

To investigate if this effect is different for diseases of different categories, we also calculate the mean *H*-score within each category and compare with the 10^4^ random networks. Fig. 3 B) shows the same effect also within categories, where the mean *H*-scores are significantly different from expected for most of the categories (see *p*-values in Tab. 3). For the non-significant categories, we find that the infectious diseases, injuries and poisonings, respiratory, and symptoms categories all seem to be more diverse in their linked diseases. For symptoms, this makes perfectly sense, as the same symptoms might co-occur with many diseases of different categories, and thus also share significant SNP-hits with co-occurring disease. Infectious diseases are only represented by five diseases in the PheNet, where two of them are located in module 2 and the rest are linked outside of the larger modules. *Chronic hepatitis* is one of the infectious diseases in module 2, which is linked to 22 other diseases of different categories. This could indicate the patients with *Chronic hepatitis* are genetically susceptible to many other type of diseases, such as *Obstructive chronic bronchitis* and *Hypoglycemia*.

**Table 3.** Mean H-score with corresponding Bonferroni adjusted p-values for each category.

| Category | PheNet |  | HUNT |  |
| --- | --- | --- | --- | --- |
| | Mean H | $p$-value | Mean H | $p$-value |
| circulatory system | 0.75 | $< 10^{-4}$ | 0.85 | $< 10^{-4}$ |
| congenital anomalies | 0.85 | $< 10^{-4}$ | 1.00 | $< 10^{-4}$ |
| dermatologic | 0.69 | $< 10^{-4}$ | 0.80 | $< 10^{-4}$ |
| digestive | 0.75 | $< 10^{-4}$ | 0.85 | $< 10^{-4}$ |
| endocrine/metabolic | 0.65 | $< 10^{-4}$ | 0.71 | $< 10^{-4}$ |
| genitourinary | 0.73 | $< 10^{-4}$ | 0.82 | $< 10^{-4}$ |
| hematopoietic | 0.58 | 0.0368 | 0.88 | 0.01 |
| infectious diseases | 0.64 | 0.2176 | 1.00 | 1 |
| injuries & poisonings | 0.81 | 0.3920 | 1.00 | 1 |
| mental disorders | 0.89 | $< 10^{-4}$ | 0.95 | $< 10^{-4}$ |
| musculoskeletal | 0.70 | $< 10^{-4}$ | 0.77 | $< 10^{-4}$ |
| neoplasms | 0.79 | $< 10^{-4}$ | 0.88 | $< 10^{-4}$ |
| neurological | 0.90 | 0.0208 | 0.93 | 0.01 |
| respiratory | 0.72 | 0.0752 | 0.76 | $< 10^{-4}$ |
| sense organs | 0.88 | $< 10^{-4}$ | 0.94 | $< 10^{-4}$ |
| symptoms | 0.54 | 1 | 0.57 | 1 |

In total, these results strongly support that most of the *H*-scores in the PheNet are higher than expected, i.e. most of the disease in the PheNet are more connected to diseases of the same category than what to be expected if they were located in a random network. Also, diseases with low *H*-scores, such as *Chronic hepatitis* and *Arthropathy*, could be interesting diseases to consider for further investigation of common genetic effects to other diseases.

### Inspecting the linkage between disease categories

Next, we consider the amount of overlap between categories to investigate if some of the categories are more linked than expected by chance. For this, we calculate the *Z*-score of the number of links connecting two categories, which is standardized against 10^4^ random simulated networks. As already concluded from the *H*-score analysis, Fig. 4 shows that most of the categories have a significant overlap with itself. In addition, the circulatory system category has significant overlap with the congenital anomalies and endocrine/metabolic categories, and the infectious diseases category has significant overlap with the dermatological and endocrine/metabolic diseases categories.

**Figure 4.**
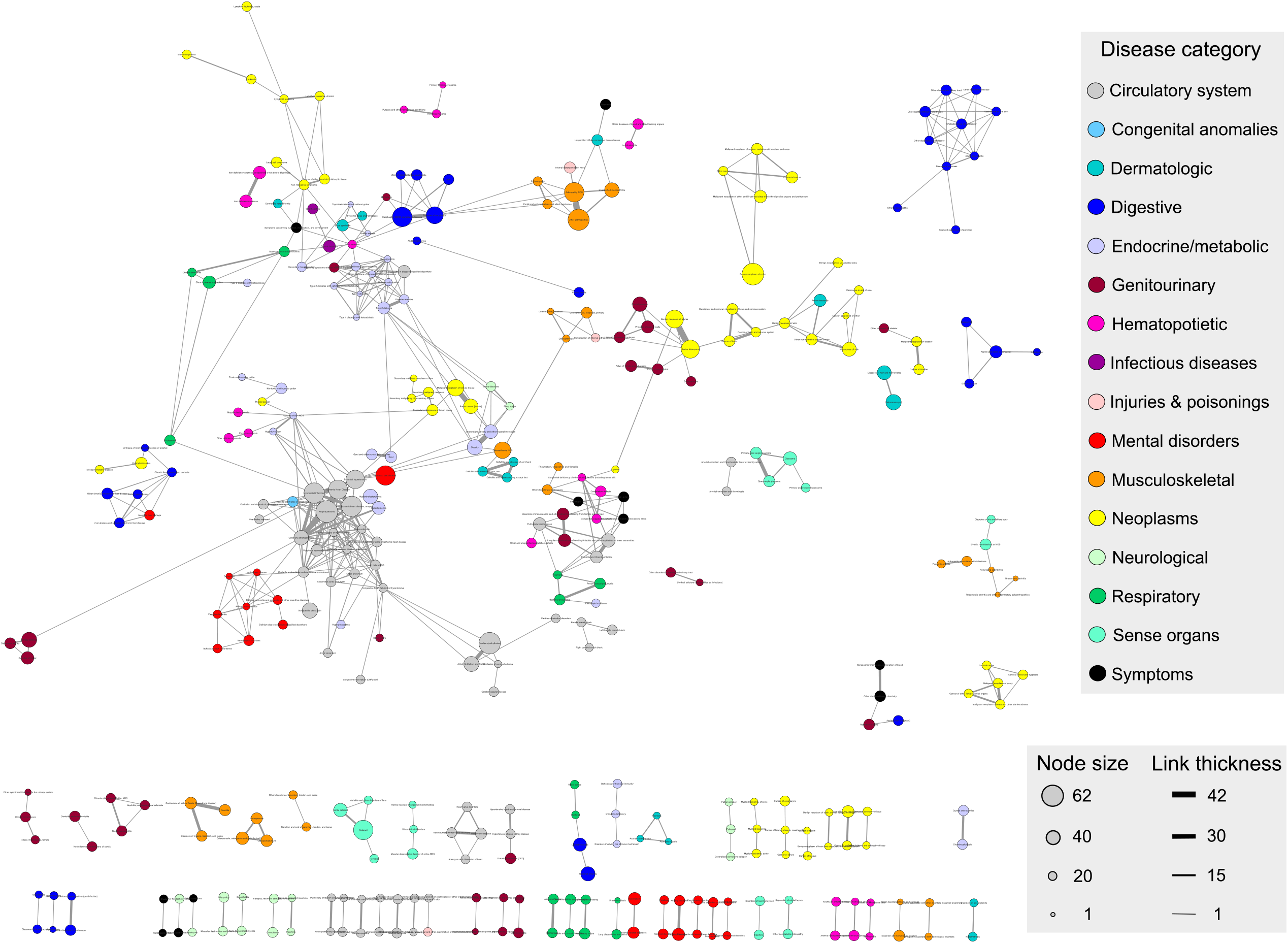
Z-score of overlap between categories. Entries are colored based on the Z-value, where Z-values corresponding to a two sided *p*-value adjusted for multiple testing (136 tests) with *p <* 0.05 are colored non-grey, and the two-sided adjusted *p*-values for these entries are shown.

Congenital anomalies are represented by five diseases in the PheNet, where two of them, *cardiac and great vessels congenital anomalies*, are linked to several circulatory system diseases in module 4. This results supports that diseases related to the cardiovascular system tend to be inherited [31, 32]. The endocrine/metabolic disease category (colored light purple in Fig. 2) is present in module 2, 3, 4 and 5, and dominates the linking between these modules. This category includes several diseases for *Type I* and *Type II diabetes*, which are known to be associated to several diseases, among them *Myocardial infarction* and *Ischemic heart disease* [36, 37] which we also observe in the PheNet. Module 2 holds two of the five infectious diseases, *Chronic and viral hepatitis*, and they are highly linked to the dermatologic and endocrine/metabolic diseases in this module.

### The HUNT sub-PheNet holds the same network properties as the PheNet

Now that we have studied the PheNet of genetically linked diseases based on UKBB study participants and its properties, we seek to investigate if these network properties are maintained when considering only disease pairs that show strong co-occurrence in the HUNT study population. Extracting the sub-network of the PheNet where disease pairs hold a Bonferroni adjusted significant *ϕ*-score, the HUNT sub-PheNet shown in Fig. 5 consists of 359 diseases with 503 links between them. This number of links is far more than expected based on simulated random networks holding the same network properties, where the mean number of significant co-occurrences is approx. 100, as shown in Fig. 6A). The HUNT sub-PheNet is hence a network showing genetically linked diseases that also show strong cooccurrences in a different study population, where the network is far denser than expected by chance.

**Figure 5.**
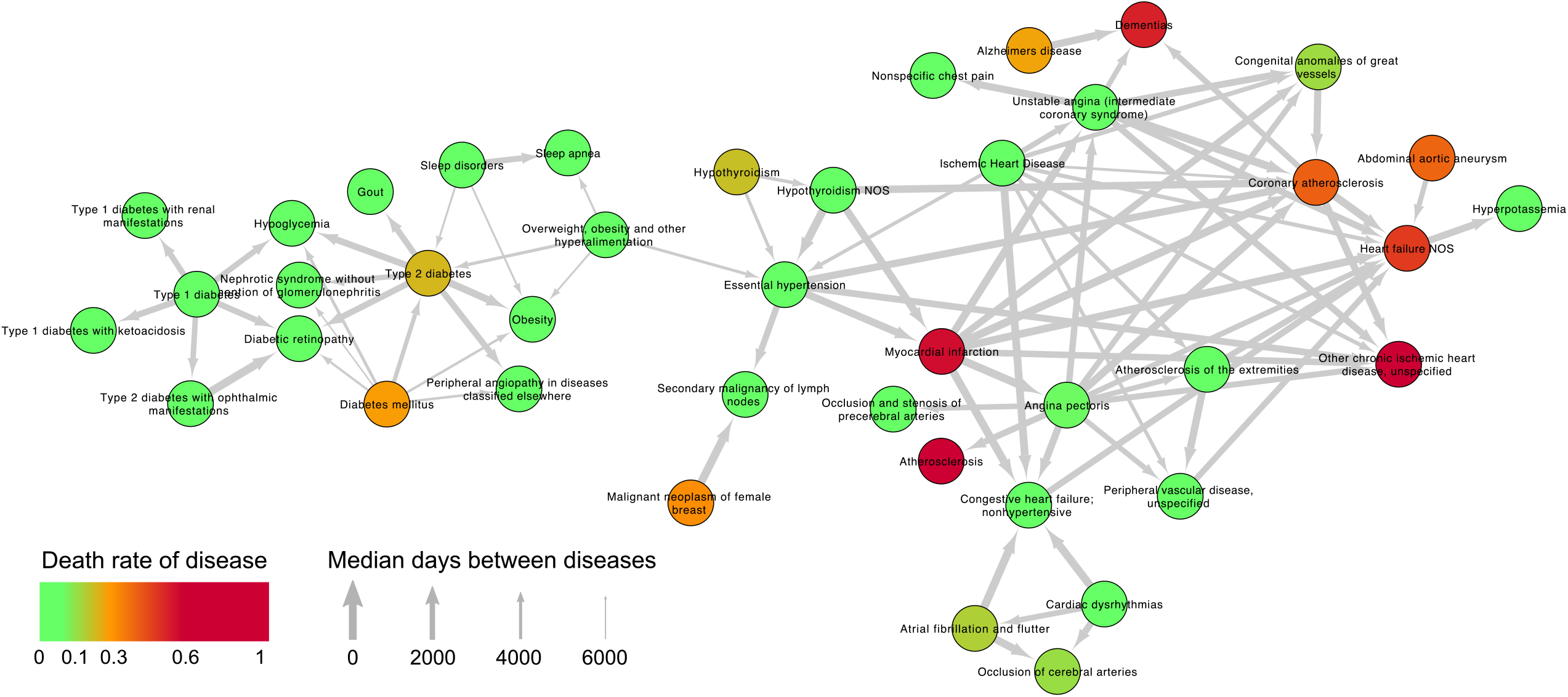
The HUNT sub-PheNet. The HUNT sub-PheNet is a sub-network of the PheNet where the only links kept are between diseases with a significant co-occurrence observed in the HUNT study. The figure features are the same as for the PheNet, and diseases with no links to other diseases (singletons) have been removed.

**Figure 6.**
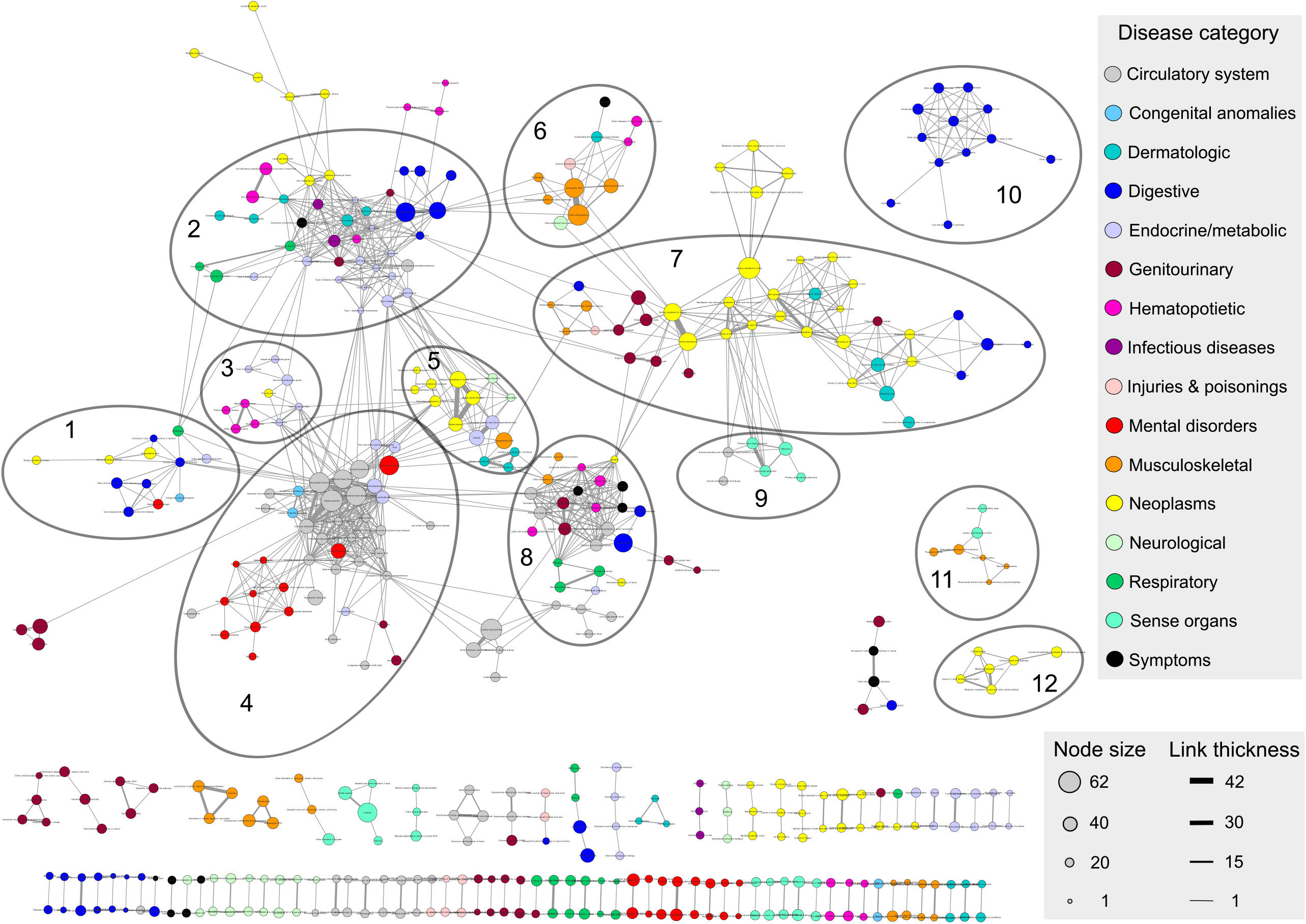
Overlap of significant unordered A) and ordered B) comorbidities. Distribution of the number of disease pairs with Bonferroni adjusted significant *ϕ*-scores in the 10^4^ random networks. The observed number of significant disease pairs from the PheNet are marked with the red arrows.

Also for the HUNT sub-PheNet, all disease categories except for pregnancy complications are represented, where the neoplasms and circulatory system categories are still the disease categories with the largest representation in the network, and congenital anomalies and infectious diseases are the categories that are the least represented in the network, as shown in Tab. 1. In general, it seems that the number of diseases represented from each category has been somewhat equally reduced prior to their presence in the PheNet, indicating that none of the disease categories stand out in terms of lacking co-occurring diseases. As a consequence of the reduced network, the mean degrees within disease categories has also been reduced. Apart from congenital anomalies with only one disease in the HUNT sub-network, who has the highest degree sharing links with 10 diseases, the circulatory system and endocrine/metabolic are still the disease categories with the highest mean degrees (5.4 and 4.2 respectably).

Even though the HUNT sub-PheNet is more sparse than the PheNet, the general structure of the network still holds when considering only pairs of diseases with strong co-occurrences. Using the *ϕ*-scores as link weights, the Louvain’s clustering method identifies 10 modules in the HUNT sub-PheNet, displaying a great overlap with the 12 modules identified in the PheNet (see Additional file 1, Fig. S2). The smallest modules from the PheNet, modules 3, 9, 11, and 12, have lost some of their links and are not included among the modules consisting of more than 5 diseases in the HUNT sub-PheNet. Modules 2 and 4 from the PheNet have been split into two separate modules in the HUNT sub-PheNet, where the group of neoplasms diseases from module 2 and the group of mental disorders from module 4 are clustered into a separate modules, named module 2a and 4b in the HUNT sub-PheNet.

Interestingly, for module 7 in the PheNet, the groups of *Colon cancer and cancer of brain* are no longer connected to a module in the HUNT sub-PheNet, showing that even though these diseases are genetically linked to the other diseases of module 7 in the PheNet, they do not show significant comorbidity in a different study population. The same goes for *Sicca syndrome* in module 2, which in the PheNet are linked to several diseases of many different categories. In the HUNT sub-PheNet, most of these links show no significant comorbidity with *Sicca syndrome*. Among the links that show significant comorbidity, we find that the links between *Obesity* and *Essential hypertension* is still present across modules, and *Essential hypertension* and *Ischemic heart disease* within module 4b.

Performing similar *H*-score analysis on the HUNT sub-PheNet, we find that the mean *H*-score within all modules and the mean *H*-score within most categories are significantly larger than expected, see Tab. 2, Tab. 3 and Additional file 1, Fig. S3. In fact, three of the four non-significant categories from the PheNet are also non-significant in the HUNT sub-PheNet, while the respiratory category seems to be less diverse in the HUNT sub-PheNet than in the PheNet. In total, these results indicating that also in the HUNT sub-PheNet based on strong co-occurrences of diseases, the diseases that are kept tend to link to diseases of the same category.

The *Z*-score analysis for the HUNT sub-PheNet (see Additional file 1, Fig. S4) shows that the only off-diagonal significant positive *Z*-score is the overlap between the circulatory system and congenital anomalies categories, as was found in the PheNet. While the significant overlap between circulatory system and endocrine/metabolic, and infectious diseases with digestive and endocrine/metabolic categories are no longer significant in the HUNT sub-PheNet, the neoplasms and circulatory system categories seem to have a smaller overlap than expected by chance. This indicates that there are few cancer diagnoses that are linked to cardiovascular diseases when considering both the genetics and the observed presence of both disease types.

### Many disease pairs show strong ordering of disease history

In the HUNT study population, we also have information regarding when the diseases occurred for each individual. Considering the date-ordered pairs of cooccurring diseases in the HUNT study population, we find that 222 of the PheNet pairs (considering both directions) show significant comorbidity, which is far more than expected based on simulations from random networks holding the same properties, see Fig. 6B). This means that a large fraction of the disease pairs observed in the PheNet are actually observed in a specific order in the HUNT study population.

Following this finding, we find that 144 disease pairs from the HUNT sub-PheNet are significantly observed with the specific ordering based on the binomial test described in the Methods section. Most of these diseases are isolated pairs or smaller groups of diseases, while 41 of them are clustered in the giant component (see Fig. 7), wheras the full network is shown in Additional file 1, Fig. S5). From the hypergeometric test of enrichment among the categories, we find that the circulatory system is over-represented among the diseases that often appear first in a pair-sequence, while the neoplasms category are under-represented among the diseases that often come last (see Additional file 2, Tab. S1).

**Figure 7.**
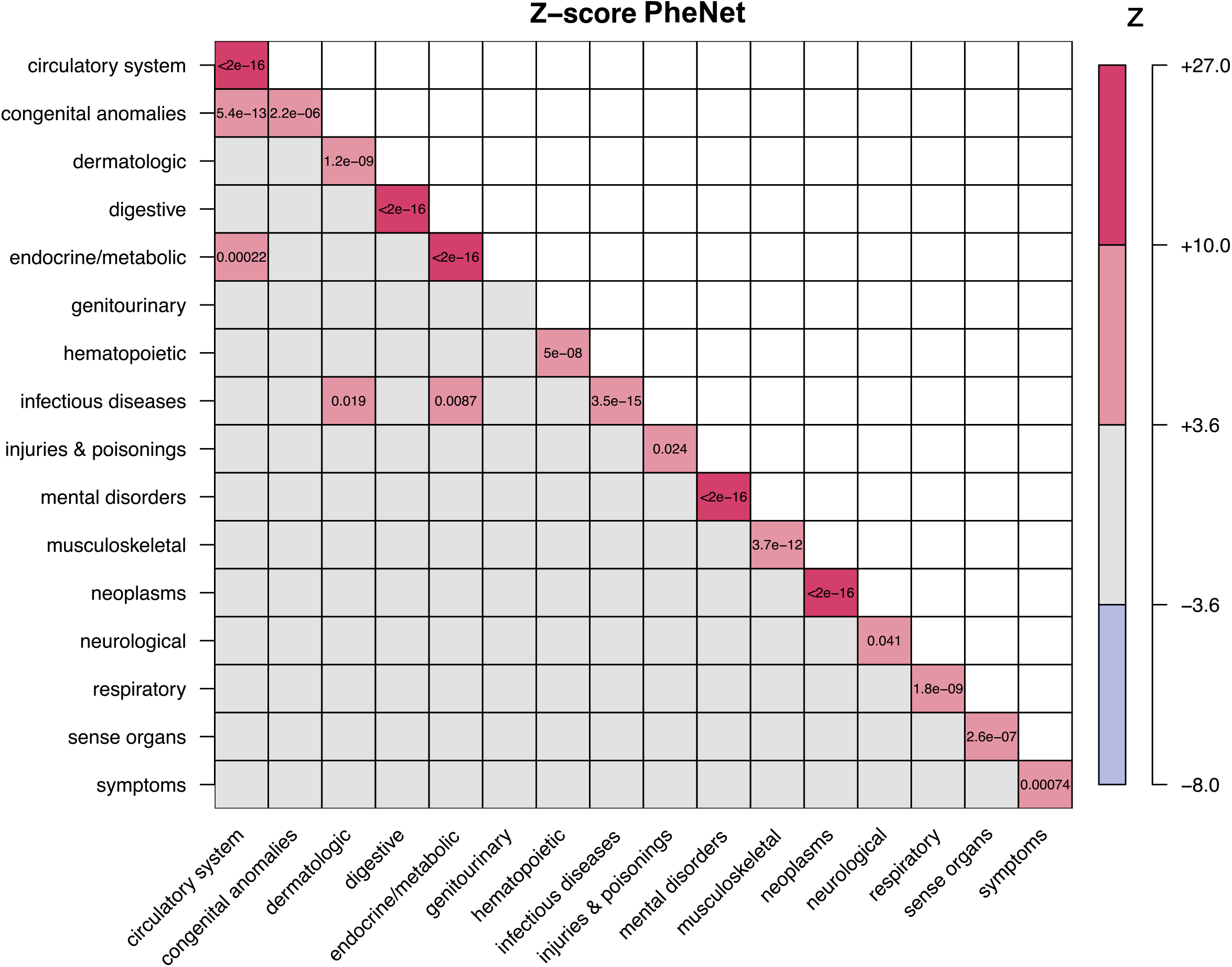
Network of ordered pairs. The giant component of ordered pairs of diseases where the arrows show the directions of the disease histories, scaled by the median time between the diagnosis. The color of the nodes represents the mortality rate of the disease.

The diseases in Fig. 7 represent mostly diabetic and cardiovascular diseases in distinct groups, where *Overweight, obesity and other hyperalimentation* connects the two groups and is more likely to precede the connected diseases. The thickness of the link represents the median time between the events (thicker means shorter time), and we observe that the time between *Overweight, obesity and other hyperalimentation* to its following diseases are much larger than the time between the cardiovascular diseases.

The diseases are colored according to their mortality rate based on the Norwegian Cause of Death registry, and we see that the groups of diabetic disorders show rater low mortality rates, where *Type 1 diabetes, Type 2 diabetes* and *Diabetes mellitus* are mostly diagnosed before the other related diseases with increasing mortality rate. It seems that the time between being diagnosed with *Type 2 diabetes* and following diseases are shorter than the time between diagnosis of *Diabetes mellitus* and the same following diseases. For the cardiovascular diseases, it appears that *Essential hypertension, Ischemic Heart Disease* and *Angina pectoris* are often diagnosed first and with low mortality rate. On the other hand, *Dementias, Heart failure NOS, Other chronic ischemic heart disease, unspecified, Atherosclerosis, Myocardial infarction* and *Coronary atherosclerosis* show high mortality rates and are often diagnosed last. The last two, *Myocardial infarction* and *Coronary atherosclerosis*, also have many outgoing links, where the time until the following events are rather short. A possible explanation could be that many patients do not die immediately after these diagnoses, but instead are unfortunate to pick up some other severe cardiovascular diseases before death due to those diseases.

## Discussion

Human disease networks offer a potential road map for both clinicians and researchers studying various forms of diseases, showing how diseases are related. Previous studies have successfully created human disease networks of genetically linked disorders, either based on diseases linked through common genes [7] or genetic information [8, 9, 10, 11, 14, 16]. Others have created disease networks entirely based on EHR-data with co-occurring diseases [26, 38]. Here, we combine the two, creating a disease network based on genomic linkage and extract the sub network of EHR-based co-occurring diseases from a different study population. With this, we show that many of the genomically linked diseases are in fact co-occurring diseases, where we overcome limitations of small sample sizes, different populations for the genetic studies, short follow-up of participants, LD-correlation, case/control imbalances and relatedness.

We are not the first to consider comorbidity among diseases based on genetically linked disease networks. Menche et. al. [39] created the interactome; genes found from OMIN and GWAS (genes with GWAS significant SNPs) linked through molecular interactions, and they showed that diseases with overlapping disease modules (overlapping genes associated to both diseases) show higher comorbidity than diseases without overlapping disease modules. However, they point out that the interactome is far from complete, and that it is biased towards much studied diseases. Park et. al. [40] focus on the overlap between diseases linked though common genes from the HDN [7] and comorbidities based on EHR-based disease histories from U.S. Medicare [26]. They show that diseases linked through common genes show higher comorbidity, where particularly diseases linked through domain-sharing genes show higher comorbidity than diseases with non-domain-sharing genes. This indicates that using SNP-linked diseases rather than gene-linked diseases could be beneficial for the study of comorbidity. Dong et. al. [14] and Sriram et. al [16] both compared SNP-based disease interactions with observed comorbidities. However, they are limited by using the same study population for both genetic testing and overlap with comorbidities. Menche et. al. [39] and Park et. al. [40] are both biased towards much studied diseases and limited by noise from translating OMIN diseases to EHR-based diseases, where the disease annotations have different nomenclatures. Their comorbidity data are strong in number of participants, but limited in the time span. The U.S. Medicare EHR-data covers only four years of disease histories for elderly patients, likely resulting in many uncovered disease co-occurrences. In our work, we do not share the strength of roughly 13 million patients, instead the HUNT study is unique in covering a total of 30 years of EHR-data for 90, 000 adult patients. With this, we argue that our SNP-based PheNet purely based on EHR data presents an unbiased and more specific disease network, and when linking to EHR-based HUNT comorbidities, we catch more of the co-occurring diseases.

Using SNP associations with diseases rather than genes, we do not consider if the SNP affects a gene that is causal for the disease. Some gene disease associations might hence be excluded as different mutations in the gene influence the expression of the gene and cause the disease. As an example, mutations in the BRCA1 and BRCA2 genes are well known to be associated with increased risk for breast cancer. However, several mutations exist, and they seem to be population specific [41]. According the the GWAS catalog, no mutations in BRCA1 and only a few mutations for BRCA2 are found to be GWAS significant. However, basing our PheNet on SNP associations, we know exactly which change in a genetic position that are associated with the disease. More than 90% of the trait-associated variants detected though GWAS are located in non-coding sequences [42], and thus, excluding all variants not coding for genes or proteins reduces the ability to study genetic causes of diseases. With current methods for functional genome annotations, one can explore the functional consequences of both coding and non-coding sequence variants detected though GWAS and PheWas [43]. Thus, genomic disease connections found in the PheNet could be used for targeted studies of functional implications of the SNPs and convergence to possible meaningful pathways, despite that the SNPs are located in non-coding regions.

Even though we argue that the networks created in this study are based on more robust methods than the works cited above, there are also some limitations. First of all, the genetically linked diseases from UKBB are purely based on white British participants of European ancestry. Hence, we cannot conclude that the diseases are genetically linked for persons with different ancestry. This is a general problem for large population genomic studies, as most are conducted with people of European ancestry, giving rising concerns about the utility of the health related outcomes from these genomic studies to patients of other ancestry and geographical locations [44].

Second, as we aimed at including as many strong disease links as possible from the PheWas data, we used no threshold for the MAF or number of participants diagnosed with the disease. This again limits the utility of the PheNet, as some diseases are linked due to SNPs that are very rare and might thus just be present for a few persons. Some diseases might also be linked where the prevalence of one or both diseases are very low. Additionally, the *p*-value threshold for inclusion of SNP disease link from the PheWas is *<* 10^*−*6^, and hence, non-GWAS significant. This choice potentially links diseases that would not have been linked with a GWAS significant threshold. However, as the main goal of this work is to identify which of the genetically linked disorders we can observe to co-occur, we argue that the questionable links will either be validated or neglected when considering only the pairs of significantly co-occurring diseases from the HUNT study.

Third, even though the HUNT data are strong in its participants rate and stretch for a very long time period, the registries used for this work are limited to *∼* 70% of the phenocodes from the UKBB, and not all of the records span the entire HUNT study time frame (1987-2017). Apart from this fact, no other population study (to our knowledge) covers hospital records for up to 30 years, which is a great advantage of our work.

Fourth, we observe that many of the diseases in our networks are quite similar. As an example, there are nine diseases corresponding to different types of diabetes. A better classification system for the diseases than the phenocodes could be beneficial for a clearer disease network. Also, some phenocodes cover the same ICD-codes, which potentially links diseases simply because they are observed to be the same diagnosis code.

Finally, as for any GWAS or PheWas, validation of the results in a separate population increases the confidence in the genomic findings. An even more robust disease network would have been one where the genomically linked diseases are validated in a separate population or with meta-analyses before extracting strongly co-occurring links. The co-occurring links could also be validated in yet another population. For the utility of these disease network in all populations, one should create disease networks based on genetic findings for all ancestries and genders, and due to genetic differences, one could also create ancestral specific disease networks. We believe that the work presented is a step towards more accurate precision medicine that, with future studies, might be beneficial for health wellness all around the world.

## Conclusions

In this study, we have created a network of genetically linked diseases that also show strong co-occurrence within a different study population. The methods for creating the PheNet overcomes limitations from previous studies, and we argue that the diseases linked in this network are based on more solid methods and datasets, and hence, being more reliable. We find that the number of overlapping disease pairs is far larger than expected by chance, and the network properties from the genetically linked disease network is mostly maintained for the subset of strong co-occurring diseases. Many diseases are connected in larger components, all disease categories, except for pregnancy complications, are included in the networks. Most diseases tend to link to other diseases of the same category, where some categories are more linked to each other than expected by chance. We have also created a directed network of consecutively occurring diseases, displaying a giant component consisting of mostly diabetic disorders and cardiovascular diseases, where the two groups are linked by following obesity and overweight. We also find that the mortality rates of these diseases are different for diseases that tend to be observed first or last.

We argue that the work presented here could be of great benefit to researchers and clinicians, and used as a resource to study and explain relationships between diseases. Hopefully, this is a step towards more precise precision medicine and we hope that further creation of such solid grounded networks for diverse ancestries could be beneficial to not only the European ancestral populations, but for people of all ancestries and genders.

## Data Availability

All data produced in the present study are available upon reasonable request to the authors

## Acknowledgements

The Trndelag Health Study (The HUNT Study) is a collaboration between HUNT Research Center (Faculty of Medicine and Health Sciences, NTNU, Norwegian University of Science and Technology), Trndelag County Council, Central Norway Regional Health Authority and the Norwegian Institute of Public Health. We want to thank clinicians and other employees at Nord-Trndelag Hospital Trust for their support and for contributing to data collection in this research project. We would like to thank the participants of the UK Biobank and the HUNT study for their contribution to research.

## Abbreviations

GWAS: Genome-wide association studies
SNP: Single Nucleotide Polymorphisms
PheWas: Phenome-wide association studies
EHR: Electronic health records
HDN: Human Disease Network
OMIN: Online Mendelian Inheritance in Man
ICD: The International Classification of Diseases
UKBB: UK Biobank
SAIGE: Scalable and Accurate Implementation of GEneralized mixed model
LD: linkage disequilibrium
HUNT: The Trndelag Health study
PheNet: Phenome-wide association network
Phenocodes: phenotype codes
PC: principal component
MAF: minor allele frequencies
HNT: Nord-Trndelag Hospital Trust (hospital records in Nord-Trndelag)
KUHR: Norway Control and Payment of Health Reimbursement
H-score: Disease homogeneity score

## Availability of data and materials

The UKBB PheWas was downloaded from https://pheweb.org/UKB-SAIGE/top_hits [45] and information regarding the study and phenotypes can be found at https://www.leelabsg.org/resources (Phenotype information table) [46]. The mapping from ICD9 and ICD10 codes are available at https://phewascatalog.org/phecodes [22, 23]. The Trndelag Health Study (HUNT) has invited persons aged 13 - 100 years to four surveys between 1984 and 2019. Comprehensive data from more than 140, 000 persons having participated at least once and biological material from 78, 000 persons are collected. The data are stored in HUNT databank and biological material in HUNT biobank. HUNT Research Centre has permission from the Norwegian Data Inspectorate to store and handle these data. The key identification in the data base is the personal identification number given to all Norwegians at birth or immigration, whilst de-identified data are sent to researchers upon approval of a research protocol by the Regional Ethical Committee and HUNT Research Centre. To protect participants? privacy, HUNT Research Centre aims to limit storage of data outside HUNT databank, and cannot deposit data in open repositories. HUNT databank has precise information on all data exported to different projects and are able to reproduce these on request. There are no restrictions regarding data export given approval of applications to HUNT Research Centre. For more information see: http://www.ntnu.edu/hunt/data.

## Ethics approval and consent to participate

Participation in the HUNT Study is based on informed consent and the study has been approved by the Data Inspectorate and the Regional Ethics Committee for Medical Research in Norway (REK: 2014/144).

## Consent for publication

Not applicable.

## Authors’ contributions

M.H. and E.A. conceived and designed research; M.H. and M.K.S analyzed data; M.H. and E.A. interpreted results of analysis; M.H. prepared figures; M.H. and E.A. drafted manuscript; M.H. and E.A. edited and revised manuscript; M.H., M.S.K. and E.A. approved final version of manuscript.

## Additional Files

Additional file 1 — Fig. S1-S5

S1: The full PheNet not reduced to include only diseases observed in the HUNT study. S2: Overlap between diseases in modules of the PheNet and the HUNT sub-PheNet. The colorbar shows the base-10 exponent of the *p*-value for the overlap. S3: Mean H-score of HUNT sub-PheNet compared to 10^4^ random networks. Mean H score across the ten largest modules A) and across the 16 phenotype categories B). The red x-es shows the results from the HUNT sub-PheNet, while the boxes with whiskers and outliers shows the results from 10^4^ simulated networks. S4: Z score of overlap between categories in the HUNT sub-PheNet. Entries are colored based on the Z-value, where Z-values corresponding to a two sided *p*-value adjusted for multiple testing (136 tests) with *p <* 0.05 are colored non-grey, and the two-sided adjusted *p*-values for these entries are shown. S5: The full network of ordered pairs of diseases where the arrows show the directions of the disease histories, scaled by the median time between the diagnosis. The color of the nodes represents the mortality rate of the disease.

Additional file 2 — Tab. S1

S1: Number of diseases that are significantly diagnosed first or last from each category. From a total of *n* such diseases (first or last), *x* are observed from each category among the *m* diseases of that category and *N − m* diseases not from that category in the HUNT sub-PheNet of *N* diseases. A hypergeometric test with variables *x, m, N − m* and *n* are used to test if the observed values are greater than expected, *P* (*X ≥ x*), or less than expected, *P* (*X ≤ x*), where the *p*-values are Bonferroni adjusted with 16 tests, where *p*-values less than 0.025 are marked with a star.

Additional file 3 — List of diseases in the PheNet (.txt)

List of diseases in the PheNet with their disease categories and in which module they are located in the PheNet and in the HUNT sub-PheNet.

