## Additional file 1 for "Phenome-wide association network demonstrates close connection with individual disease trajectories from the HUNT study"

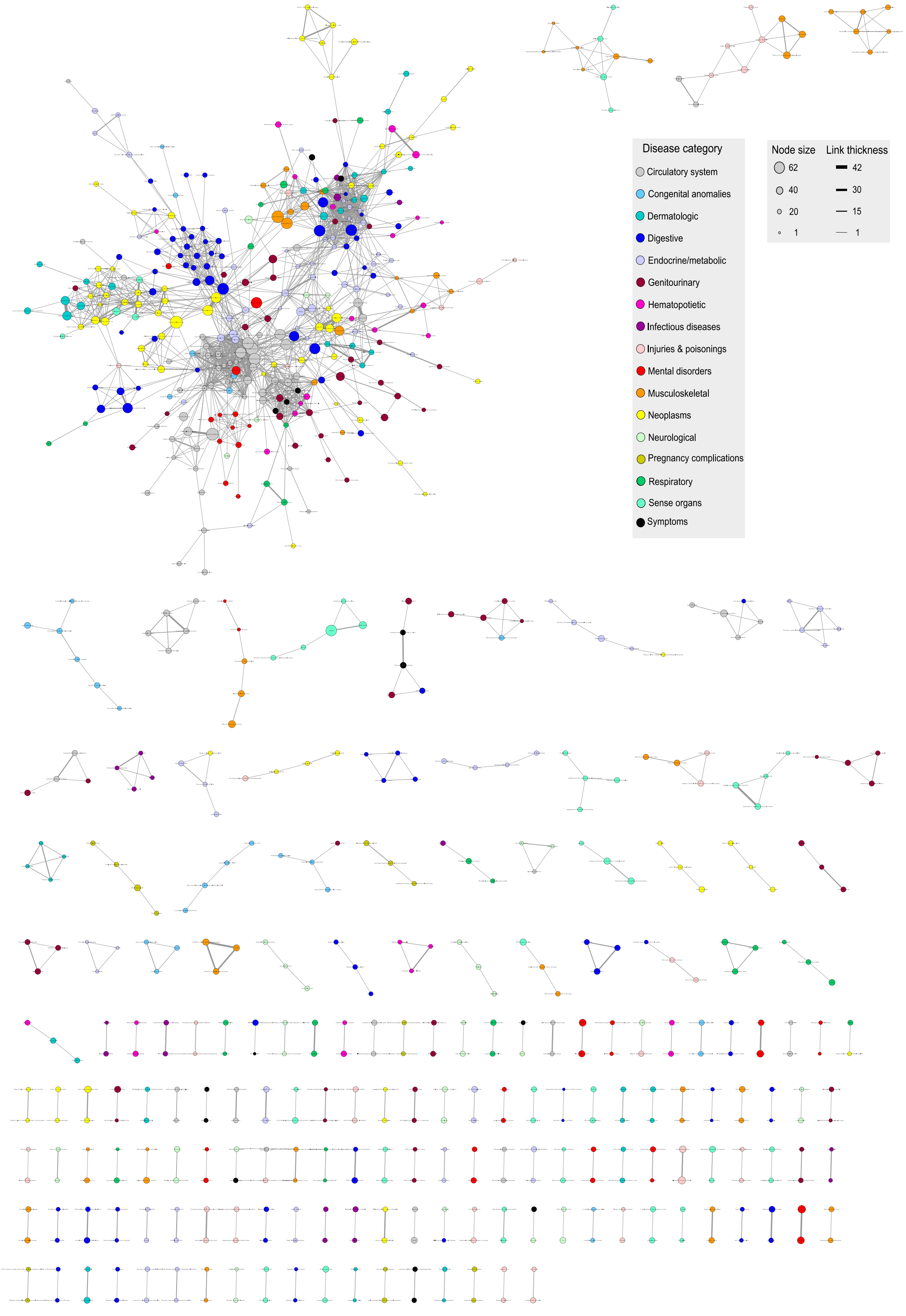


Fig. S1: The full PheNet not reduced to include only diseases observed in the HUNT study

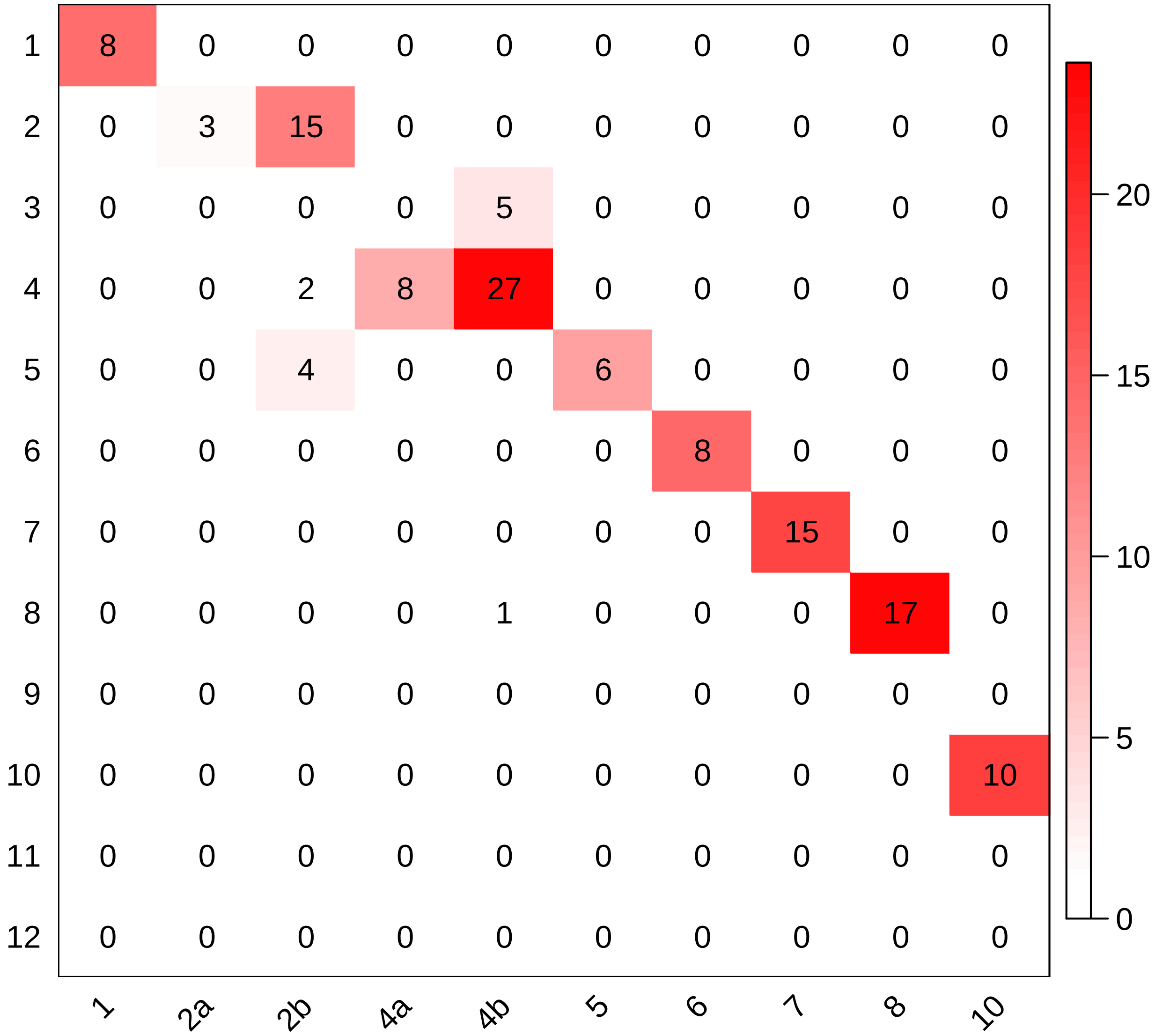


Fig. S2: Overlap between diseases in modules of the PheNet and the HUNT sub-PheNet. The colorbar shows the base-10 exponent of the $p$-value for the overlap.

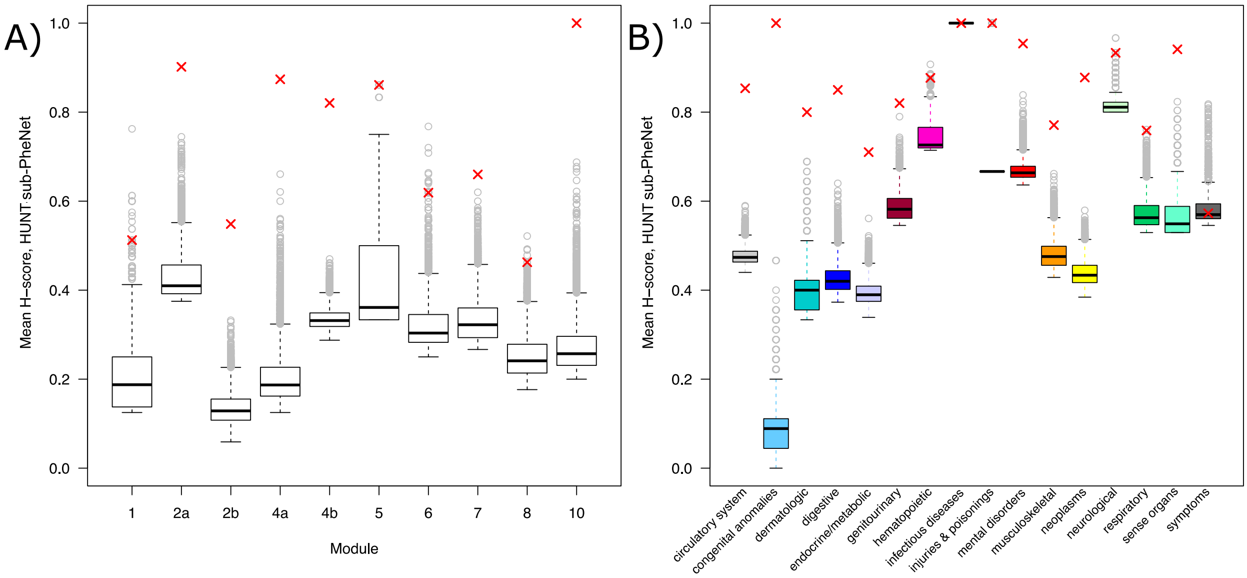


Fig. S3: Mean H-score of HUNT sub-PheNet compared to $10^4$ random networks. Mean H score across the ten largest modules A) and across the $16$ phenotype categories B). The red x-es shows the results from the HUNT sub-PheNet, while the boxes with whiskers and outliers shows the results from $10^4$ simulated networks.

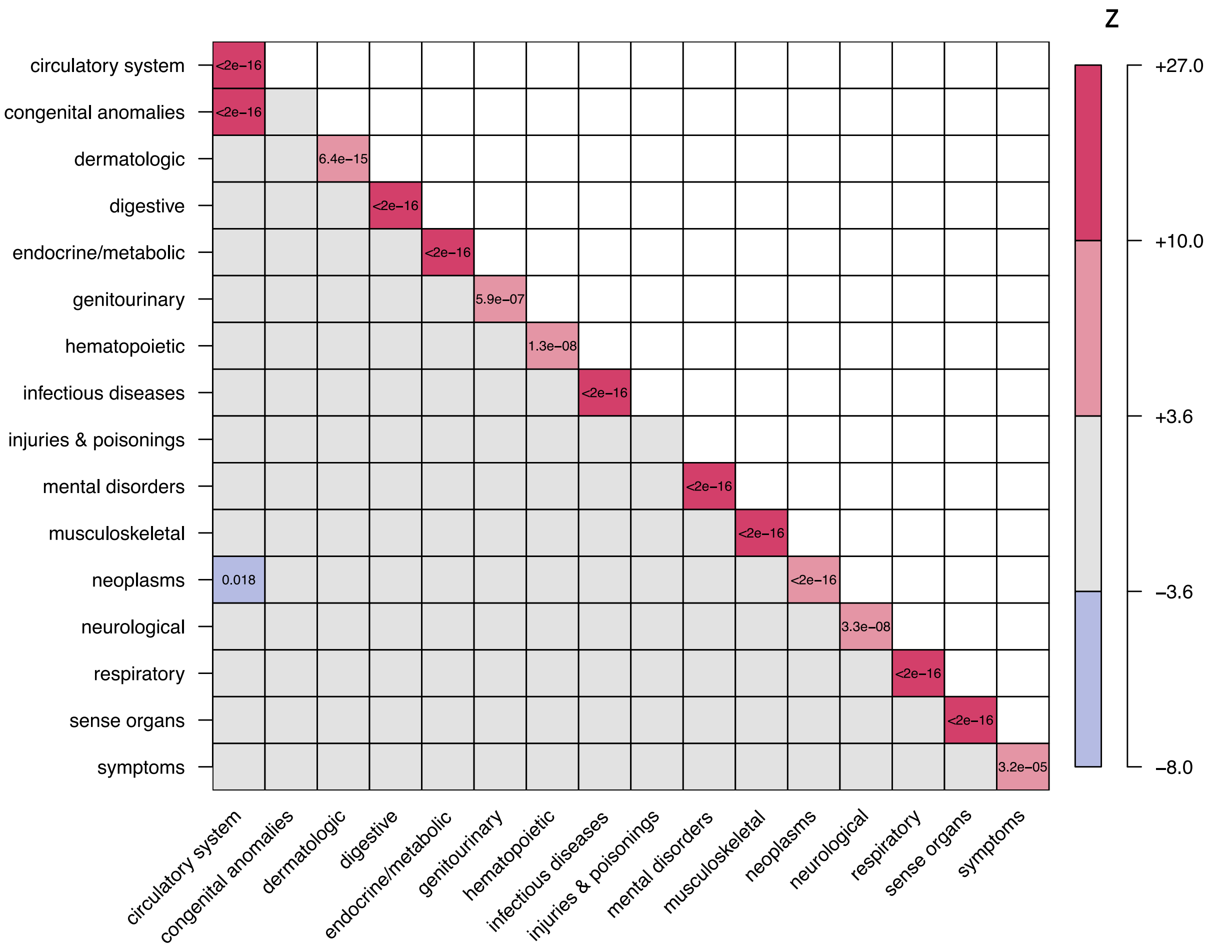


Fig. S4: Z score of overlap between categories in the HUNT sub-PheNet. Entries are colored based on the Z-value, where Z-values corresponding to a two sided $p$-value adjusted for multiple testing (136 tests) with $p < 0.05$ are colored non-grey, and the two-sided adjusted $p$-values for these entries are shown.

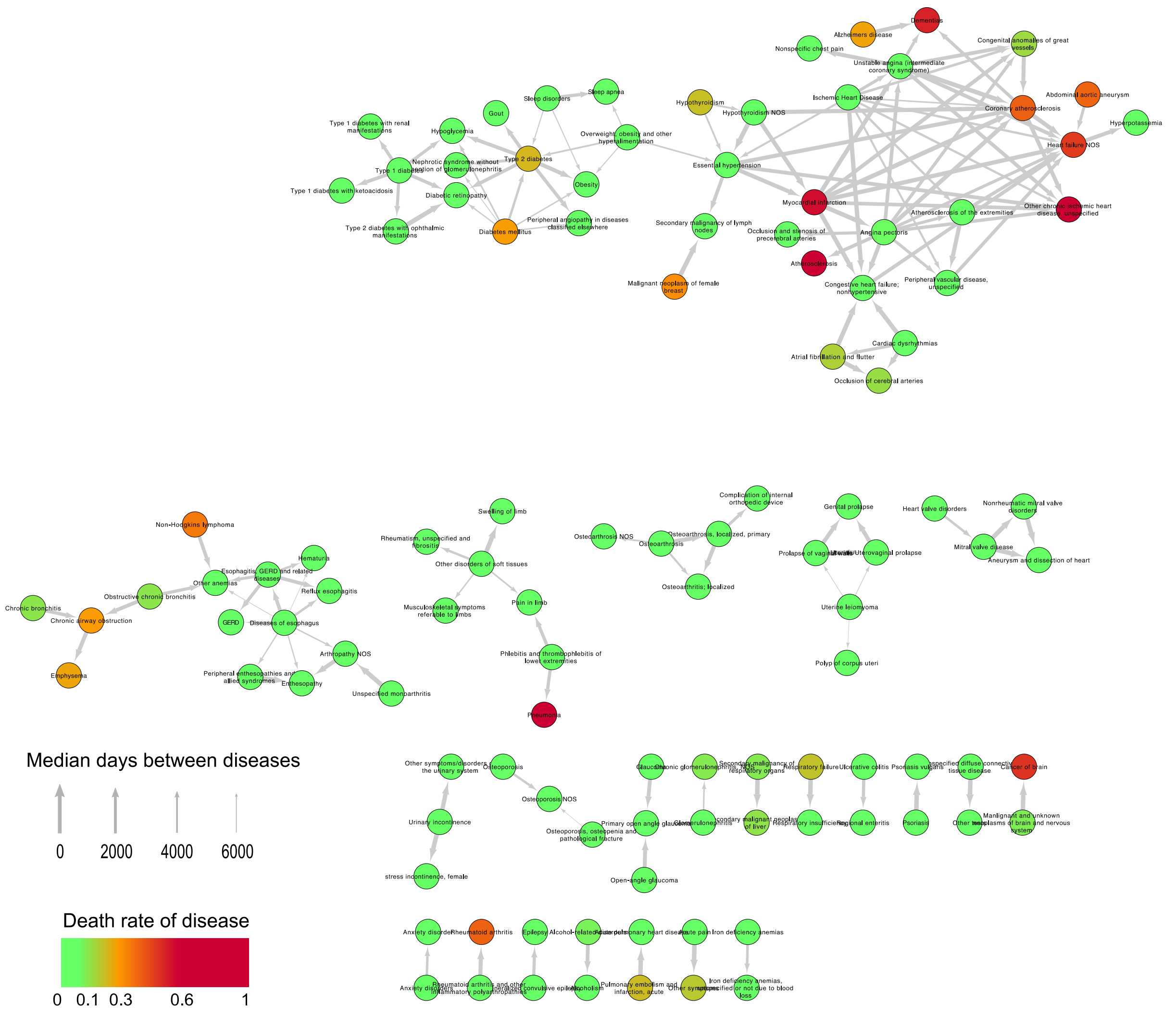


Fig. S5: The full network of ordered pairs of diseases where the arrows show the directions of the disease histories, scaled by the median time between the diagnosis. The color of the nodes represents the mortality rate of the disease.
