## Additional file 2 for "Phenome-wide association network demonstrates close connection with individual disease trajectories from the HUNT study"

Tab. S1: Number of diseases that are significantly diagnosed first or last from each category. From a total of $n$ such diseases (first or last), $x$ are observed from each category among the $m$ diseases of that category and $N-m$ diseases not from that category in the HUNT sub-PheNet of $N$ diseases. A hypergeometric test with variables $x, m, N-m$ and $n$ are used to test if the observed values are greater than expected, $P(X \geq x)$, or less than expected, $P(X \leq x)$, where the $p$-values are Bonferroni adjusted with 16 tests, where $p$-values less than $0.025$ are marked with a star.

| | Diseases diagnosed first | | | | | Diseases diagnosed last | | | | |
|---|---|---|---|---|---|---|---|---|---|---|
| Category | $x$ | $m$ | $N-m$ | $P(X \geq x)$ | $P(X \leq x)$ | $x$ | $m$ | $N-m$ | $P(X \geq x)$ | $P(X \leq x)$ |
| circulatory system | 18 | 54 | 305 | 0.0214* | 1 | 20 | 54 | 305 | 0.0355 | 1 |
| congenital anomalies | 1 | 1 | 358 | 1 | 1 | 1 | 1 | 358 | 1 | 1 |
| dermatologic | 2 | 15 | 344 | 1 | 1 | 1 | 15 | 344 | 1 | 1 |
| digestive | 3 | 35 | 324 | 1 | 1 | 4 | 35 | 324 | 1 | 1 |
| endocrine/metabolic | 7 | 30 | 329 | 1 | 1 | 10 | 30 | 329 | 1 | 1 |
| genitourinary | 4 | 33 | 326 | 1 | 1 | 9 | 33 | 326 | 1 | 1 |
| hematopoietic | 1 | 21 | 338 | 1 | 1 | 2 | 21 | 338 | 1 | 1 |
| infectious diseases | 0 | 2 | 357 | 1 | 1 | 0 | 2 | 357 | 1 | 1 |
| injuries & poisonings | 0 | 3 | 356 | 1 | 1 | 1 | 3 | 356 | 1 | 1 |
| mental disorders | 3 | 22 | 337 | 1 | 1 | 3 | 22 | 337 | 1 | 1 |
| musculoskeletal | 9 | 28 | 331 | 0.5521 | 1 | 9 | 28 | 331 | 1 | 1 |
| neoplasms | 5 | 55 | 304 | 1 | 0.8688 | 3 | 55 | 304 | 1 | 0.0122* |
| neurological | 3 | 15 | 344 | 1 | 1 | 2 | 15 | 344 | 1 | 1 |
| respiratory | 4 | 17 | 342 | 1 | 1 | 4 | 17 | 342 | 1 | 1 |
| sense organs | 2 | 17 | 342 | 1 | 1 | 1 | 17 | 342 | 1 | 1 |
| symptoms | 0 | 11 | 348 | 1 | 1 | 5 | 11 | 348 | 0.9061 | 1 |
| $n$ | 62 | | | | | 75 | | | | |
| $N$ | | 359 | | | | | 359 | | | |
